# Multiplexed Isothermal Nucleic Acid Detection Using Sequence-Specific Cleavage Mediated by Repair Endonucleases

**DOI:** 10.64898/2026.05.29.26354475

**Authors:** Padric M. Garden, Yudan Li, Vel Murugan, Alexander A. Green

## Abstract

Rapid, portable detection of multiple nucleic acid targets is essential for infectious disease surveillance and precision oncology. CRISPR-based diagnostics have set a high bar for sensitivity and single-base specificity, yet their reliance on collateral nuclease activity complicates multiplexing, integration with amplification, and point-of-care deployment. Here we present TIMBER (Templated Incision Mediated By Endonucleases of Repair), a non-CRISPR, isothermal platform that achieves comparable performance without nonspecific nuclease activity. TIMBER uses a repair endonuclease to cleave probes containing abasic sites only when they are hybridized to a matched target. The system exhibits strong specificity, enabling single-nucleotide polymorphism discrimination. TIMBER provides an analytical limit of detection 12 pM without preamplification or 1 copy per µL (1.6 aM) with preamplification through RT-PCR or RT-LAMP, with results observable by visual fluorescence or on lateral flow. We deploy TIMBER to detect SARS-CoV-2 in clinical saliva samples and further demonstrate multiplex detection of four targets enabling identification of *EGFR* mutations in lung cancer samples. This approach offers a flexible, rapid, and low-instrument ready solution for diverse nucleic acid diagnostics, from viral detection to cancer genotyping.

**Graphical Abstract:** 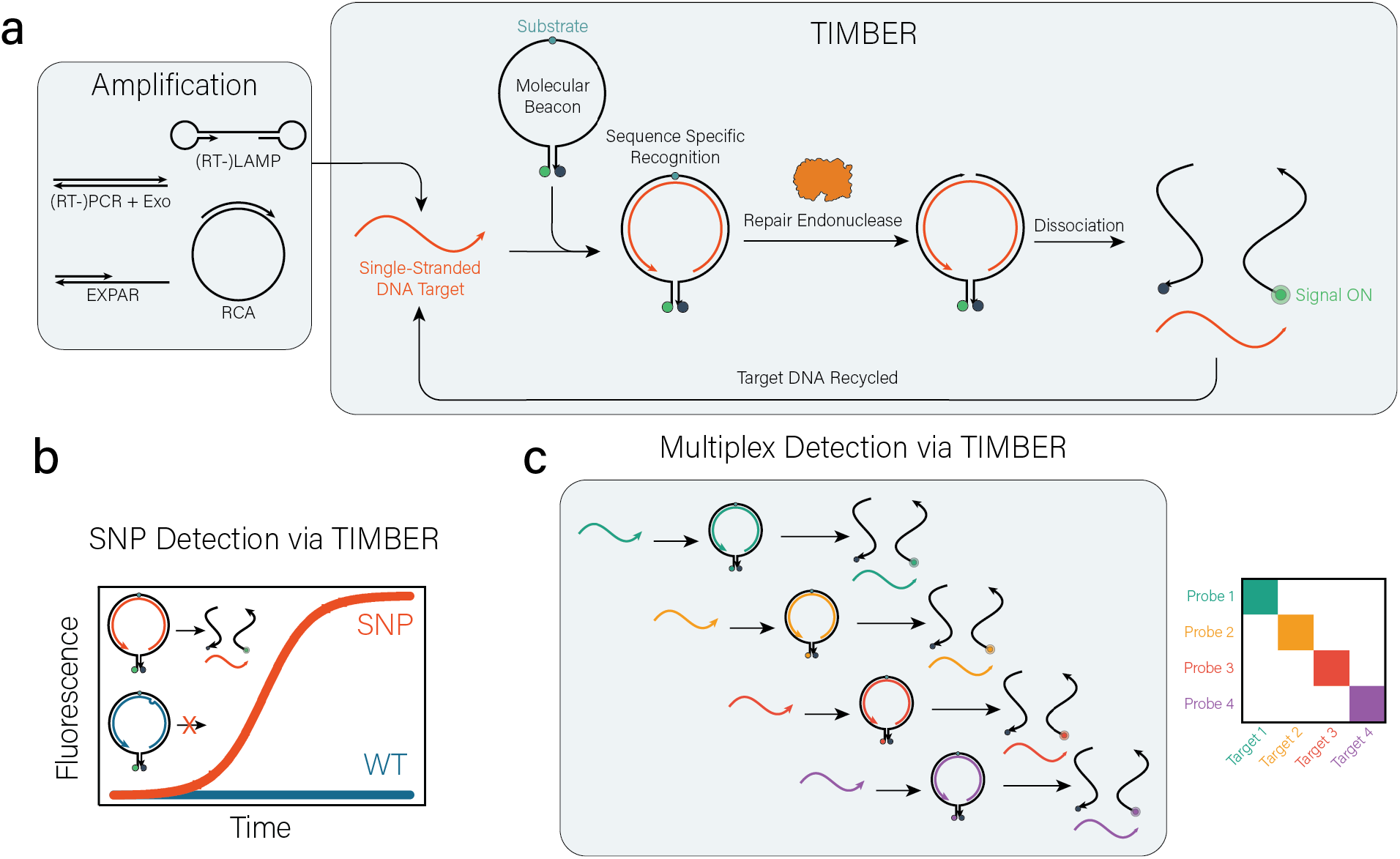

## INTRODUCTION

Nucleic acid-based diagnostics enable highly specific detection of many infectious diseases, disease-related biomarkers, and the classification of mutations and variants.^1,2^ For point-of-care use, these diagnostics often utilize isothermal amplification methods, which lower instrumentation needs by avoiding thermal cycling and provide faster reaction speeds. A variety of isothermal amplification methods have been developed, such as recombinase polymerase amplification (RPA), helicase-dependent amplification (HDA), loop-mediated isothermal amplification (LAMP), rolling circle amplification (RCA), and exponential amplification reaction (EXPAR).^1–3^ These amplification techniques are often paired with detection strategies which can mitigate nonspecific amplification, reducing the likelihood of false-positive results, and provide an easily detectable readout, potentially enabling low-instrumentation detection.^1,2^

CRISPR-associated (Cas) enzymes have proven to be powerful detection strategies for use in nucleic acid diagnostics.^1^ In particular the inducible collateral cleavage activity of Cas12, Cas13 and Cas14 enable highly sensitive, highly specific detection of nucleic acids when paired with amplification approaches like RPA or LAMP.^4–6^ Despite their many advantages, these approaches are limited due to the relatively indiscriminate nature of collateral cleavage, which reduces multiplexing capacity and compatibility with other assay components, and they often rely on the use of multiple Cas enzymes with carefully designed probe sequences,^7^ toehold-mediated strand displacement (TMSD) DNA circuitry,^8^ or the spatial separation of reactions.^9,10^ As an alternative, Argonaute-based sensing systems utilize guide oligonucleotides to specifically recognize and cleave target nucleic acids, offering the distinct advantage of functioning without PAM sequence constraints.^11^ However, these systems are currently limited by slower reaction speeds, a reliance on thermophilic enzymes that require high operational temperatures, and the potential for non-specific cleavage in the absence of guide DNA.^11,12^ Non-enzymatic sensors based on structural switching of nucleic acids, such as toehold switches^13^ and aptaswitches^14^, can provide highly specific^15,16^, molecular logic^17^, and multiplex detection^14^ in point of care formats. However, the sensitivity of these systems is in the nanomolar range for visible readout, necessitating the use of isothermal amplification when used against clinical samples.^18,19^

DNA repair endonucleases are enzymes that target substrates that are indicative of DNA damage, such as abasic sites (AP Sites), nonstandard bases (e.g. deoxyuridine, deoxyinosine, 8-Oxoguanine), or mismatches. Their repair function occurs through cleavage of the damaged strand.^20,21^ In nature, these enzymes play key roles in the recognition and excision of damaged bases for base excision repair pathways.^20,22^ Moreover, repair endonucleases have attractive properties for biotechnology applications due to their substrate specificity – they often recognize one damaged or modified base within a sequence of DNA – as well as their ability to interface with polymerases and other DNA modifying enzymes through the generation of nicks. For example, repair endonucleases have previously been used to control the initiation of RCA through cleavable primers,^23^ to release DNAzymes in the presence of a DNA target,^24^ and to generate a fluorescent readout for LAMP reactions.^25^

Here we report the development of a multiplexable, isothermal readout method for nucleic acid detection utilizing repair endonucleases, which we have named Templated Incision Mediated By Endonucleases of Repair (TIMBER) which can be paired with a variety of amplification techniques including (RT-)PCR, (RT-)LAMP, EXPAR, and RCA. Using *Thermus thermophilus* endonuclease IV in conjunction with a fluorogenic probe containing a 1’,2’-dideoxyribose abasic site, we demonstrate a limit of detection down to 12 pM of target DNA when TIMBER is used alone and down to 1 copy per µL (1.6 aM) of target DNA or RNA when TIMBER is coupled with preamplification. TIMBER has strong sequence specificity and careful probe design enables discrimination of single-nucleotide polymorphisms (SNPs). Moreover, unlike approaches based on CRISPR collateral cleavage, this approach cleaves probes in a sequence specific manner, enabling easy multiplex detection of up to 4 targets through the use of orthogonal probes with a single enzyme. We also demonstrate the suitability of TIMBER to low-instrument fluorescent and lateral flow readouts. Lastly, we apply the multiplex and SNP detection capabilities of TIMBER toward detecting clinically relevant *EGFR* mutation in lung cancer samples validated via qPCR.

## RESULTS

### Development of TIMBER Assays

TIMBER relies on the capacity of suitable repair endonucleases to preferentially cleave nonstandard bases in double-stranded DNA (dsDNA), while leaving these same modified bases unscathed in single-stranded DNA (ssDNA) form (Fig. 1a). Briefly, a probe can be engineered to contain the substrate sequence recognized by a repair endonuclease, embedded within a region complementary to the target sequence of interest. In the absence of the target, the substrate sequence is single-stranded and not discerned by the endonuclease. However, upon hybridization of the probe to its target DNA, the repair endonuclease recognizes the substrate sequence in double-stranded form and cleave the probe without affecting the target DNA. This selective cleavage allows the target DNA to remain intact and participate in subsequent rounds of probe binding and cleavage, enabling signal amplification.

**Figure 1.**
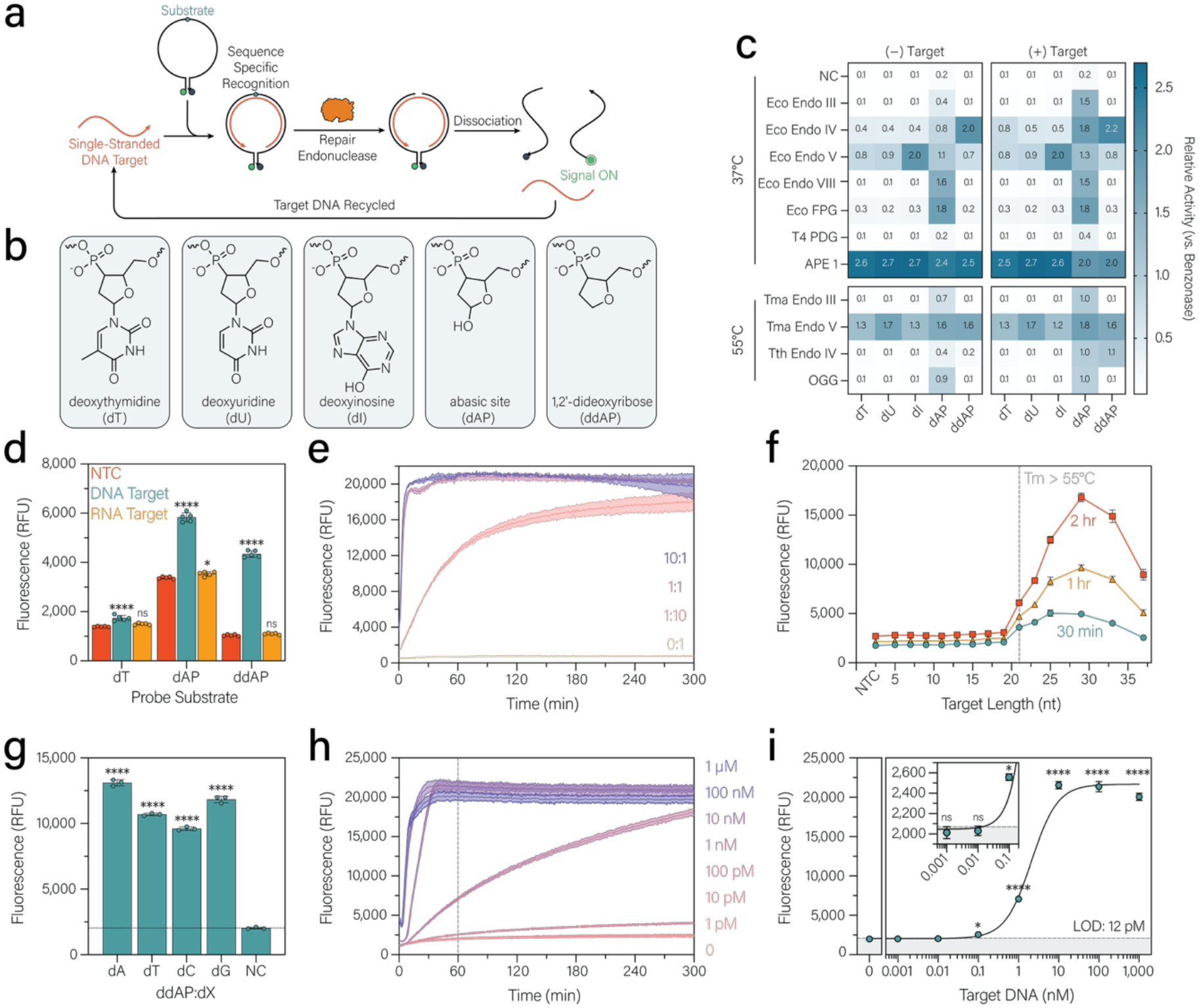
Testing and optimization of DNA repair endonucleases for TIMBER. **a**, Schematic of TIMBER detection scheme. This molecular beacon probe contains a substrate for repair endonucleases and hybridizes to a single-stranded DNA target, enabling the repair endonuclease to nick the probe. The resulting complex dissociates, generating a fluorescent signal and allowing the probe to be recycled for additional cleavage events. **b**, Structures of substrates examined in this study, including dU, dI, dAP, and ddAP, with dT serving as a control. **c**, Comparison of repair endonucleases, showing their activity on the probe alone versus in the presence of the complementary target DNA. Enzyme activities are reported relative to the nonspecific nuclease benzonase. **d**, Response of the Tth endonuclease IV-based TIMBER assay on probes containing dT, dAP, or ddAP in the presence of complementary DNA and RNA targets. **e**, Kinetic fluorescence profile of TIMBER at various ratios of target DNA to probe. **f**, Fluorescence response of TIMBER assays to target DNA lengths ranging from 5 to 37 nt, measured at 30 minutes, 1 hour, and 2 hours. The vertical line indicates the target length with a melting temperature ≥55°C, which corresponds to the reaction temperature used. **g**, Fluorescence response of TIMBER to targets containing each standard base opposite the abasic site in the probe. **h**, Kinetic fluorescence profile of TIMBER in response to a 27-nt target at concentrations ranging from 1 pM to 1 µM. The vertical line indicates the 1-hour time point used for the calibration curve in i. **i**, Calibration curve of TIMBER against an arbitrary target DNA, showing an analytical limit of detection of 12 pM.

We thus sought to evaluate a broad range of repair endonucleases for their ability to differentiate between nonstandard bases in ssDNA and dsDNA. We first incubated each enzyme with a molecular beacon fluorescence-quencher (FQ) probe containing various substrates recognized by repair endonucleases: deoxyuridine (dU), deoxyinosine (dI), abasic site (dAP), and 1’,2’-dideoxyribose (ddAP), along with deoxythymidine (dT) as a control (Fig. 1b, Supplementary Table 1). These probes were tested in the presence or absence of a complementary 25-nt target DNA. To account for varying assay temperatures, fluorescence measurements were normalized to the same FQ probe conditions pre-incubated with the indiscriminate nuclease benzonase (Fig. 1c). Under these conditions, Tth endonuclease IV exhibited a strong preference for dsDNA when used with a dAP or ddAP-containing FQ probe.

We then examined Tth endonuclease IV in greater detail. We first optimized assay buffer conditions, as well as enzyme and probe concentrations (Supplementary Fig. 1). Probes containing dT, dAP, and ddAP were tested with DNA and RNA complements (Fig. 1d). Tth endonuclease IV appears to have no activity on RNA:DNA hybrids under these conditions. Increased leakage was observed when using the dAP substrate, which is likely due to the natural propensity of abasic sites to undergo spontaneous strand scission, leading to high background fluorescence in the absence of a target. In contrast, the synthetic ddAP substrate enabled enzyme-driven cleavage while minimizing background signal. We then examined TIMBER under both excess target and excess probe conditions, demonstrating that the system approaches the same maximal fluorescence, (Fig. 1e) indicating that TIMBER is a multiturnover system.

We next sought to characterize TIMBER-based nucleic acid detection. We determined the optimal target hybridization length to between 25 and 33 nt, and no activation was seen when the T_m_ was less than the reaction temperature (55°C) (Fig. 1f, Supplementary Table 2). We then examined the activation of TIMBER by targets containing each possible base across from the ddAP site (Fig. 1g). The system was strongly activated regardless of the base with some preference towards dA and dG. Lastly, we examined the sensitivity of TIMBER against a synthetic 29-nt target. We successfully detected targets down to 100 pM and calculated an analytical limit of detection of 12 pM for an assay time of 1 hour (Fig. 1h-i). This limit-of-detection was calculated from a 1/Y^2^ - weighted 4PL curve as the concentration corresponding to 3 standard deviations above the negative mean. We also implemented a TIMBER assay with the mesophilic enzyme *E. coli* endonuclease III (Supplementary Fig. 2).

### Implementing TIMBER Assays with SNP Specificity

We evaluated the specificity of TIMBER assays by testing targets containing one to three mismatches at various positions along a 25-nt sequence (Fig. 2a-c, Supplementary Table 3). Targets with two or three mismatches within eight nucleotides of the abasic site produced minimal signal, highlighting the strong sequence specificity of TIMBER (Fig. 2b-c). Although single mismatches were tolerated at some positions, we identified a region three to five nucleotides upstream of the abasic site where a single mismatch inhibited cleavage, suggesting these positions may be useful for SNP discrimination.

**Figure 2.**
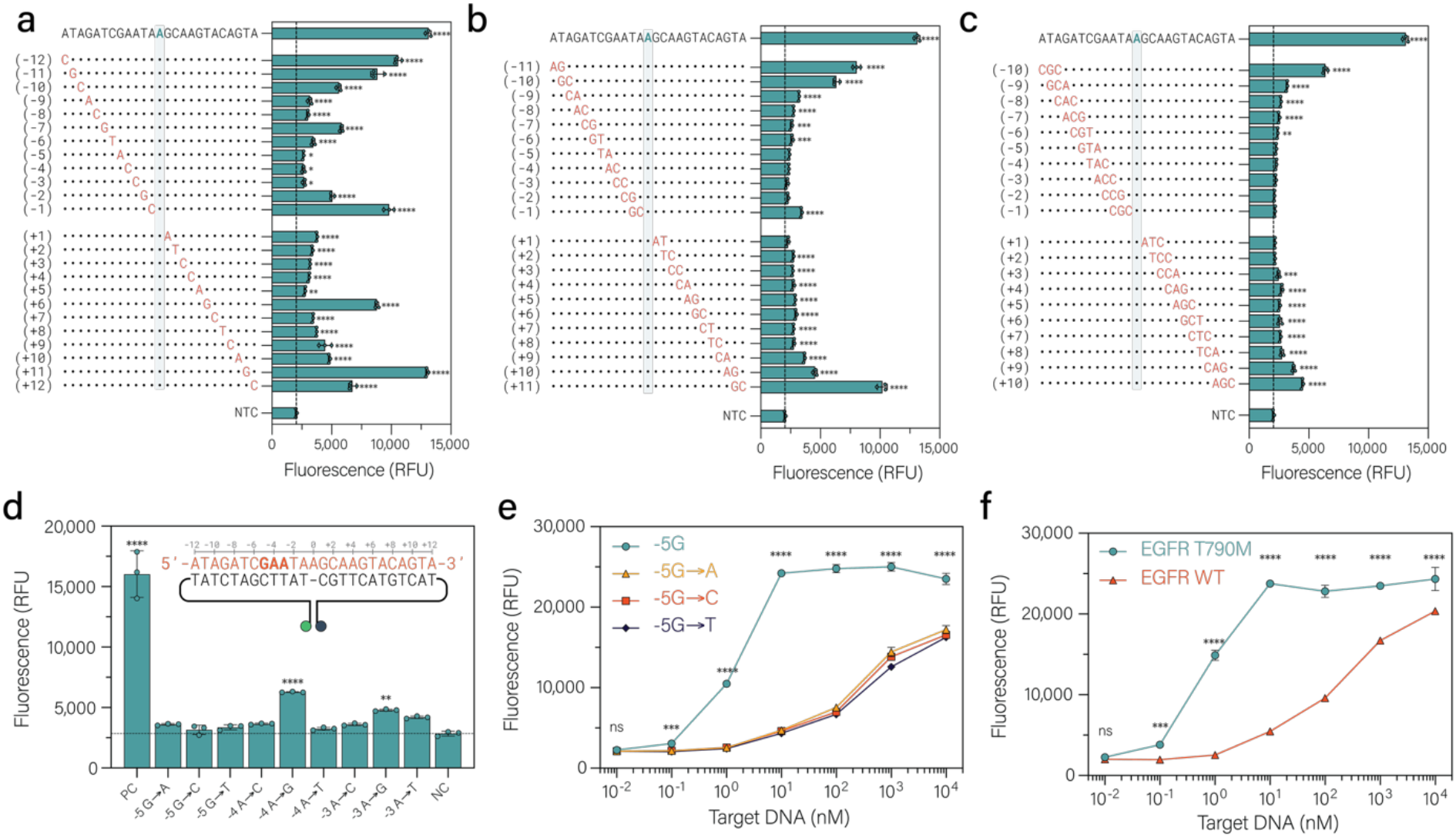
Evaluation of the specificity of TIMBER for SNP detection. **a-c**, Fluorescence response of TIMBER to single mismatches (a), two consecutive mismatches (b), and three consecutive mismatches (c) along the target DNA. Shaded region marks the position corresponding to the abasic site in the probe. **d**, Response of TIMBER to each possible mismatch in the −5, −4, and −3 positions relative to the abasic site. **e**, Sensitivity of TIMBER towards a specific target compared to the 3 possible mismatches in the −5 position of an arbitrary target. **f**, Sensitivity of TIMBER targeting *EGFR* WT versus T790M SNP.

To further explore this specificity, we introduced all possible single-nucleotide polymorphisms (SNPs) within that critical region (Fig. 2d). Notably, any mismatch at the −5 position significantly reduced activation, indicating that probes can be optimized for SNP detection by positioning the abasic site five nucleotides downstream of the SNP. We next compared the system’s response to its target versus SNP-containing variants (Fig. 2e). Although some activation was observed above 10 nM, the assay consistently distinguished the matching target from SNP-containing sequences at the −5 position. Given that most nucleic acid amplification methods yield DNA concentrations in the hundreds of nanomolar range, the observed specificity should be sufficient for SNP discrimination when coupled with a pre-amplification step.

We then confirmed that TIMBER could detect a biologically relevant SNP. In this process, we found that care needs to be taken regarding the probe’s secondary structure to ensure that the abasic site is not within a stem under the assay conditions as this leads to nonspecific probe cleavage (Supplementary Fig. 3). A probe was designed against *EGFR* T790M by placing the SNP in the −5-position relative to the abasic site. This probe was examined against a synthetic target containing a 50-nt region around the SNP and the equivalent region of wild-type (WT) DNA, demonstrating similar discrimination as seen with the arbitrary target (Fig. 2f).

### Integration of TIMBER with Isothermal Amplification Techniques

We next investigated whether TIMBER could be effectively combined with nucleic acid amplification methods to enhance sensitivity and expand its applicability to mRNA and miRNA targets (Fig. 3a). As TIMBER assays require single-stranded DNA targets, we selected amplification techniques capable of producing single-stranded amplicons. For mRNA detection, we demonstrated that RT-PCR using one primer protected with phosphorothioate modifications and another containing a 5′ phosphate could generate single-stranded DNA after treatment with lambda exonuclease. Additionally, RT-LAMP could be employed by targeting TIMBER probes to single-stranded loop regions within the LAMP product. Using these methods, we achieved sensitive detection down to 1 copy per µL (1.6 aM) for human *ACTB* mRNA (Fig. 3b) and the SARS-CoV-2 *E* gene (Fig. 3c).

**Figure 3.**
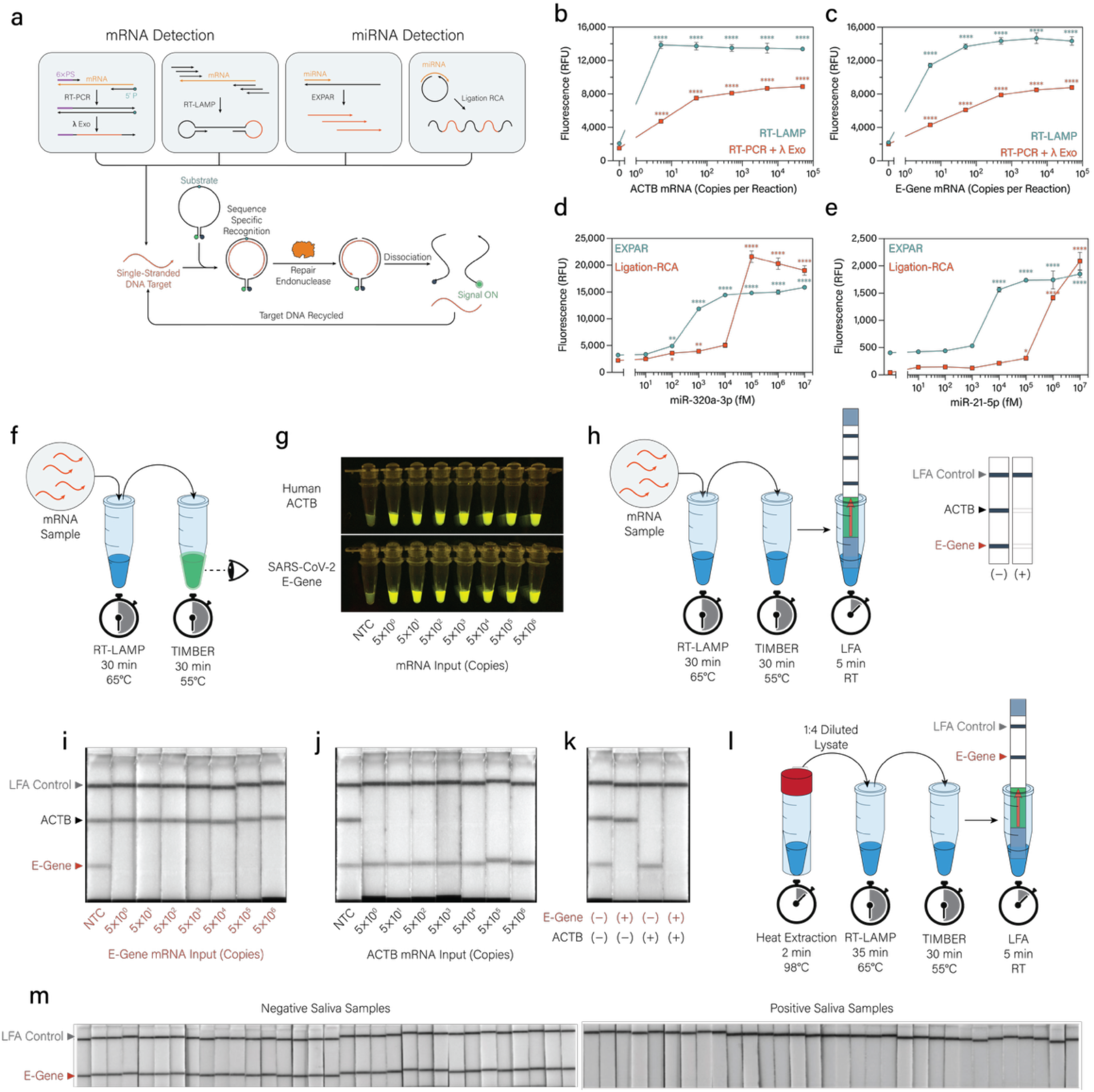
Integration of TIMBER with a variety of amplification and readout methodologies. **a**, Schematic illustrating how different pre-amplification approaches can be integrated with TIMBER for mRNA and miRNA detection. For mRNAs, RT-PCR products generated using one phosphorothioate-protected primer and one 5′-phosphorylated primer that can be treated with lambda exonuclease to yield ssDNA, which is then targeted by TIMBER probes. RT-LAMP produces single-stranded loop regions similarly accessible to TIMBER probes. For miRNAs, EXPAR and ligation-based RCA each generate ssDNA products that can be detected by TIMBER. **b**, Detection of human ACTB mRNA using TIMBER coupled with RT-PCR or RT-LAMP. **c**, Detection of the SARS-CoV-2 *E* gene using TIMBER coupled with RT-PCR or RT-LAMP. **d**, Detection of miR-320a-3p using TIMBER coupled with EXPAR or ligation-based RCA. **e**, Detection of miR-21-5p using TIMBER coupled with RT-PCR or RT-LAMP. **f**, Schematic of how RT-LAMP and TIMBER can be combined for a visual fluorescence readout. A sample is incubated at 65 °C for 30 minutes in an RT-LAMP reaction, then transferred to the TIMBER reaction at 55 °C for 30 minutes before visual inspection. **g**, Fluorescence output of the RT-LAMP-TIMBER approach for *ACTB* and the *E* gene. **h**, Schematic of a duplex lateral flow assay (LFA) readout for TIMBER. Following RT-LAMP at 65 °C for 30 minutes, the product is transferred to the TIMBER reaction and incubated at 55 °C for 30 minutes before adding LFA running buffer and the LFA strip, which reveals the result within five minutes. The lower test line corresponds to the *E* gene probe, and the upper test line corresponds to the *ACTB* probe. The absence of a band indicates a positive result. **i**, Duplex RT-LAMP-TIMBER LFA response for the SARS-CoV-2 *E* gene. **j**, Duplex RT-LAMP-TIMBER LFA response for *ACTB*. **k**, Orthogonal detection of the *E* gene and *ACTB* using a duplex LFA. **l**, Saliva sample processing workflow (left) with LFA readout from 32 SARS-CoV-2-positive saliva samples and 32 negative saliva samples after heat extraction, RT-LAMP, and TIMBER assay (right).

To extend the method to miRNA targets, we employed EXPAR (Exponential Amplification Reaction) and ligation-based RCA (Rolling Circle Amplification). EXPAR directly generates single-stranded products readily targeted by TIMBER probes, while ligation-based RCA produces extended single-stranded DNA containing numerous copies of the target sequence. Using these amplification strategies, we successfully detected miR-320a-5p at concentrations as low as 100 fM with EXPAR and 100 pM with ligation-based RCA (Fig. 3d). Similarly, detection limits of 10 pM and 1 nM were achieved for miR-21 using EXPAR and ligation-based RCA (Fig. 3e).

### Suitability of TIMBER for Low-Resource Readouts

To assess TIMBER’s suitability for low-resource settings, we first evaluated its capacity for a visual fluorescent readout. Notably, both TIMBER and RT-LAMP reaction require only isothermal amplification and can generate a visual signal within an hour (Fig. 3f). Using this approach, we detected both human *ACTB* mRNA and the SARS-CoV-2 *E* gene at concentrations as low as 1 copy per µL (1.6 aM) from a 5-µL sample (Fig. 3g).

Next, to enable multiplex detection with minimal instrumentation, we adapted TIMBER to a lateral flow assay (LFA) format. This was achieved using universal duplex lateral flow strips with test lines for biotin and digoxigenin, along with gold nanoparticles targeting FAM. Specifically, a probe with 5′ FAM and 3′ biotin was used for the SARS-CoV-2 *E* gene, while a probe with 5′ FAM and 3′ digoxigenin was used for human *ACTB*. Because TIMBER cleaves these probes, the LFA operates in an inverted format, where the absence of a test line indicates the presence of target (Fig. 3h). Using this approach, as few as five copies of SARS-CoV-2 *E* gene (Fig. 3i) and human *ACTB* gene (Fig. 3j) were detected. Moreover, we demonstrated orthogonal detection of each of these targets (Fig. 3k).

To validate the clinical utility of TIMBER, we then evaluated its performance on human saliva samples for SARS-CoV-2 detection. A panel of 64 saliva samples, comprising 32 confirmed SARS-CoV-2 negatives and 32 SARS-CoV-2 positives were tested. Saliva/viral mRNA was extracted through a rapid 2-minute heat treatment at 98°C directly from thawed human saliva. The heat-treated samples were then diluted 1:4 into water and subjected to RT-LAMP. TIMBER and LFA readout were then performed with a 5′ FAM and 3′ biotin labeled probe. In these clinical samples, TIMBER demonstrated 100% sensitivity and 100% specificity for detecting the SARS-CoV-2 *E* gene (Fig. 3l).

### Multiplex Detection of *EGFR* Mutations in Lung Cancer Samples

We then sought to validate TIMBER’s multiplexing and SNP detection capability in human samples. In lung cancer, the mutational profile of the epidermal growth factor receptor (*EGFR*) gene has proven a useful tool in determining the optimal treatment regime. Exon 19 deletions (Ex19del) are among the most common mutations, and can be indicative of a cancer that will respond to *EGFR* Tyrosine Kinase Inhibitors (TKIs), similarly the common resistance mutation T790M (typically the result of a c.2369C>T SNP) can be indicative of poor response to some TKIs and that Osimertinib should be utilized.^26,27^ Accordingly, we implemented a four-plex TIMBER panel targeting the wild-type and T790M mutant *EGFR*, along with a highly conserved Exon 19 reference site and a region of Exon 19 commonly missing in Ex19del mutants (Fig. 4a).

**Figure 4.**
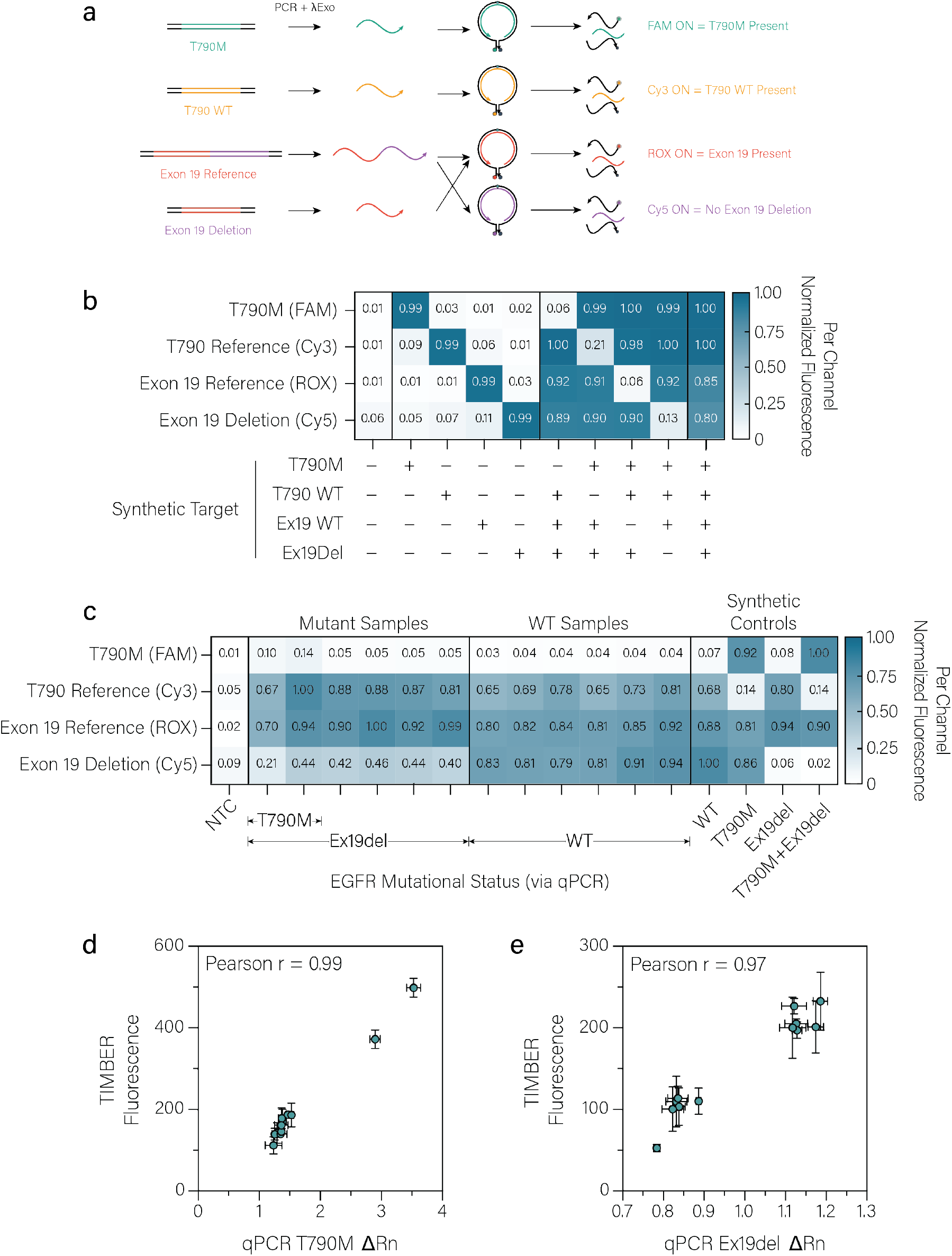
Multiplex detection of *EGFR* mutations in lung cancer samples. **a**, Schematic demonstrating how four-plex TIMBER can be applied to *EGFR* mutational analysis. Two-plex PCR is run using primers targeting the regions near the T790M SNP and in an area of Exon19 that contains a highly conserved region, and a region commonly deleted in lung cancer. Lambda exonuclease converts the products to a single stranded form. The resulting product is added to a four-plex TIMBER reaction containing probes for T790M, T790 WT, Exon 19 Reference, and Exon 19 Deletion, enabling a readout of the mutational status of *EGFR*. **b**, Four-plex TIMBER detection of synthetic targets for TIMBER probes targeting *EGFR* T790 WT, T790M SNP, Exon 19 Reference and Exon 19 Deletion. **c**, Four-plex TIMBER detection of 12 samples, 6 with known *EGFR* mutations, and 6 wild-type samples as well as 4 synthetic controls corresponding to each possible mutational status. **d**, Correlation of TIMBER fluorescence and qPCR ΔRn for T790M. **e**, Correlation of TIMBER fluorescence and qPCR ΔRn for Exon 19 deletion.

First, we first confirmed that these four probes provided orthogonal detection of their target sequence when using synthetic oligonucleotides (Fig. 4b). We then examined PCR amplicons of genomic DNA samples collected from six lung cancer samples with known *EGFR* mutations and six wild-type samples, as well as amplicons of synthetic DNA with each possible mutational profile using four-plex TIMBER (Fig. 4c). The mutational status showed good agreement between the two techniques, with a Spearman’s correlation of 0.99 and 0.97 for T790M and Ex19del respectively (Fig. 4d, e) indicating that TIMBER can accurately determine the mutational status of *EGFR*.

## DISCUSSION

In this study, we introduced TIMBER (Templated Incision Mediated By Endonucleases of Repair) and demonstrated its capacity for highly specific, isothermal detection of nucleic acids. By leveraging the sensitivity of Tth endonuclease IV to mismatches near the abasic site in the probe, we achieved high sequence specificity, thus enabling straightforward multiplex detection and efficient SNP discrimination. When coupled with RT-PCR or RT-LAMP, TIMBER is capable of detecting as few as 5 copies (1 copy/µL) of a target mRNA. Moreover, by coupling TIMBER with diverse amplification approaches, including (RT-)LAMP, (RT-)PCR, EXPAR, and ligation-based RCA, we extended its applicability to a wide range of nucleic acid targets, such as genomic DNA, mRNA and miRNAs, outlining the method’s broad utility. Overall, these findings underscore TIMBER’s potential as a versatile platform capable of ultrasensitive and highly specific multiplex detection of nucleic acids in a variety of contexts, from low-instrument viral diagnostics to genomic SNP profiling.

Placed alongside other nucleic acid detection methods, particularly CRISPR-based assays and traditional repair endonuclease approaches, TIMBER provides a distinct advantage in achieving sequence-specific cleavage without collateral nuclease activity while maintaining similar sensitivity. Without amplification, the sensitivity of TIMBER is similar to that of many CRISPR-based approaches, and when coupled with RT-LAMP our detection limit rivals that of the most sensitive amplification-coupled CRISPR techniques.^1,7^ Collateral cleavage leads to severe limitations in how many CRISPR assays can be applied, as it can destroy assay components required for amplification, and adds complexity in multiplexing, often requiring multiple enzymes or carefully designed probe sequences.^7,8,28^ Our results seem to reinforce previous demonstrations of Tth endonuclease IV ability to cleave DNA probes in a double-stranded specific manner^25^ and its sensitivity to SNPs within close proximity of the abasic site^29^, while further expanding characterizing the enzyme’s activity and expanding its use to more pre-amplification techniques.

Nonetheless, TIMBER’s utility is constrained by its requirement for single-stranded DNA as input, a notable limitation when compared to systems like Cas12a, which can be activated directly by double-stranded DNA. This necessitates additional steps during sample preparation or amplification to generate single-stranded targets, potentially complicating the workflow. Furthermore, the design of TIMBER probes demands careful attention to secondary structure; for instance, positioning the abasic site within a stem region can lead to nonspecific cleavage. These challenges underscore the importance of meticulous probe design and optimization to ensure reliable performance, while also highlighting potential avenues for future improvements.

Unexpectedly, we observed that Tth endonuclease IV exhibited markedly improved discrimination between double-stranded and single-stranded DNA when used in buffer conditions not recommended by the manufacturer (50 mM NaCl, 10 mM Tris-HCl, 10 mM MgCl_2_, 100 µg/mL Recombinant Albumin, pH 7.9) compared to the manufacturer’s recommended buffer (20 mM Tris-HCl, 10 mM (NH_4_)_2_SO_4_, 10 mM KCl, 2 mM MgSO_4_, 0.1% Triton X-100, pH 8.8). Moreover, experiments with Eco endonuclease III revealed the potential for double-stranded-specific cleavage of ddAP when NaCl concentrations were increased to approximately 200 mM. These observations indicate that reaction buffer conditions can drastically alter the activity of repair endonucleases, suggesting that systematic optimization of buffer composition could further enhance TIMBER performance.

The potential for using mesothermic enzymes such as Eco endonuclease III also presents an exciting opportunity for TIMBER to function at lower temperatures (near 37°C). Transitioning to a mesothermal assay could significantly enhance the platform’s utility, particularly for point-of-care diagnostics where low-instrumentation setups and lower temperature amplification techniques like RCA or RPA are advantageous.

Future work will involve a systematic evaluation of additional enzymes, buffer conditions, probe designs, and the integration of TIMBER into one-pot formats with isothermal amplification techniques. These efforts will help establish an expanded TIMBER toolbox for diverse diagnostic and research applications.

## MATERIALS AND METHODS

### Materials

RNA and DNA oligos were purchased from Integrated DNA Technologies (IDT) or Genscript. Eco Endonuclease III, Eco Endonuclease IV, Eco Endonuclease V, Eco Endonuclease VIII, Eco FPG, T4 PDG, APE-1, Tma Endonuclease III, Tth Endonuclease IV, OGG, Antarctic Thermolabile UDG, NEBuffer r1.1, NEBuffer r2.1, NEBuffer r3.1, rCutSmart, ThermoPol, Isothermal Amplification Buffer I, Isothermal Amplification Buffer II, WarmStart Multi-Purpose LAMP/RT-LAMP 2X Master Mix (with UDG), and Q5 High-Fidelity 2X Master Mix were purchased from New England Biolabs (NEB). Tma Endonuclease V was purchased from ThermoFisher Scientific. HybriDetect Universal Lateral Flow Assay Kits were purchased from Milenia Biotec. AmpliScribe T7 High Yield Transcription (IVT) Kits were purchased from Biosearch Technologies. Lung cancer genomic DNA was purchased from BioChain. *EGFR* T790M SNP genotyping assay and TaqPath ProAmp Master Mix were purchased from ThermoFisher Scientific. Monarch Spin RNA Cleanup Kits were purchased from NEB.

### Screening Repair Endonucleases

Molecular beacon probes labeled at the 5′ end with FAM and at the 3′ end with Black Hole Quencher-1 (BHQ-1), and containing deoxyuridine, deoxyinosine, 1′,2′-dideoxyribose, or deoxythymidine were purchased from IDT. To generate a natural abasic site, the deoxyuridine-containing probe (1 µM) was treated with Antarctic Thermolabile UDG (0.02 U/µL) in 1X NEBuffer r2.1 for 30 min at 30 °C. Each 10-µL reaction contained 100 nM of probe, 0.25 µL of enzyme, and 1 nM of target DNA (for positive samples) in 1X NEBuffer r2.1. For positive reference controls for each probe, the enzymes were replaced with the indiscriminate nuclease Benzonase (Millipore Sigma, 0.05 U/µL); fluorescence readings were then normalized to these controls to account for variation among probes. Reactions were incubated at either 37°C (for mesophilic enzymes) or 55°C (for thermophilic enzymes) for 4 h. Probe and target sequences used in the screening are listed in Supplementary Tables 1 and 2.

### TIMBER Assays

Unless otherwise noted, TIMBER assay reaction mixtures consisted of 100 nM of each ddAP-containing probe, 0.2 U/µL Tth Endonuclease IV (NEB), and 1 µL of target in 1X NEBuffer r2.1, with a total reaction volume of 10 µL for each replicate. Reactions were sealed to prevent evaporation and monitored at 55°C using a BioTek Synergy Neo2 plate reader (Agilent). Single-plex FAM-based TIMBER assays were recorded using the following conditions: Excitation: 484±20, Emission: 530±25, Bottom read, Gain: 100. Unless otherwise indicated, reported fluorescence values are the results at one hour.

### In Vitro Transcription

AmpliScribe T7 High Yield Transcription Kit was used following the manufacturer’s instructions using DNA templates containing the desired sequence on a T7 promoter. Reaction mixes were incubated at 37°C for 2 hours. 1 µL RNase-Free DNase I was added, and the reaction was incubated at 37°C for 15 minutes. The resulting RNA was purified using a Monarch Spin RNA Cleanup Kit and quantified using its A260 on a Thermo Scientific™ NanoDrop™ One.

### LAMP/RT-LAMP Sample Preparation

RT-LAMP reactions were prepared according to the manufacturer’s instructions (NEB, M1708S) in a total volume of 25 µL using a full set of 6 LAMP primers (FIP and BIP: 1.6 µM each, F3 and B3: 0.2 µM each, LF and LB: 0.4 µM each) and using 5 µL of sample. Reactions were incubated at 65°C for 30 minutes. Duplex RT-LAMP reactions for *ACTB* and *E* gene were performed by adding both sets of primers each at the same concentrations. All LAMP/RT-LAMP primer and TIMBER probe sequences are listed in Supplementary Table 4.

### PCR/RT-PCR-Exonuclease Sample Preparation

RT-PCR reactions were prepared according to the manufacturer’s instructions (NEB, M3029S) in a total volume of 25 µL, using 400 nM of each primer. Forward primers were modified with six phosphorothioate bonds at their 5′ ends, while reverse primers included a 5′ phosphate modification. All PCR/ q-PCR primer sequences are listed in Supplementary Table 5.

PCR was carried out on an Applied Biosystems ProFlex Thermocycler with the following cycling conditions: 25°C for 30 seconds; 55°C for 10 minutes; 95°C for 1 minute; followed by 50 cycles of 95°C for 10 seconds and 60°C for 1 minute. After completion of RT-PCR, 1 µL of lambda exonuclease (NEB) was added, and reactions were incubated at 37°C for 30 minutes to generate single-stranded product prior to addition to TIMBER assays. For multiplex *EGFR* mutational analysis, two sets of primers targeting the T790 region and the Exon 19 region were used, with all four primers each at 400 nM.

### EXPAR Sample Preparation

EXPAR reactions were performed by adding 1 µL of miRNA sample to a 10 µL reaction containing 10 nM DNA Template, 0.1 U/µL Nt.BstNBI, 0.3 U/µL Bst 3.0 Pol, 10 ng/µL ET SSB, 1 U/µL Murine RNase Inhibitor, 200 µM dNTPs in 1X NEBuffer r3.1. The reactions were incubated at 55°C for 20 minutes.

### Ligation-RCA Sample Preparation

In the ligation-RCA reactions,1.5 µL of padlock probe (1 µM) and 1.5 µL of miRNA target (various concentrations) were first annealed with 0.3 µL of splintR buffer (NEB, M0375S) at 80°C for 5 minutes then gradually cooled down to room temperature. Padlock probes were 5′-phosphorylated ssDNA probes that hybridize to miRNA targets via their 5′ and 3′ termini. Then a 15 µL of ligation-RCA reactions were prepared by mixing the following components: 3.3 µL of annealing product, 0.8 µL of splintR ligase (NEB, M0375S), 1.2 µL of SplintR buffer (NEB, M0375S), 0.2 µL of RCA enzyme (Cytiva, 410478), 5 µL of RCA reaction buffer (Cytiva, 410482) and 4.5 µL of nuclease free water. Reactions were incubated at 30°C for 90 minutes. RCA padlock probe sequences are listed in Supplementary Table 4.1, TIMBER probe sequences for miRNA detection are listed in Supplementary Table 4.2.

### Visual Fluorescence Readout

Visual readout TIMBER reactions consisted of 200 nM ddAP-containing probes, 0.2 U/µL Tth Endonuclease IV (NEB), and 4 µL of target in 1X NEBuffer r2.1, with a total reaction volume of 40 µL for each sample. After incubation at 55°C for 30 minutes, samples were placed on a blue transilluminator and covered with an orange filter. Samples were then imaged using Adobe Lightroom on an iPhone 12 mini using an exposure time of 1 second and an ISO of 40.

### Lateral Flow Readout

Lateral flow readout TIMBER probes were modified to remove the hairpin stem, and end-labeled with FAM and biotin for the SARS-CoV-2 *E* gene probe and with FAM and digoxigenin for the *ACTB* reference probe. Reactions consisted of 1 nM each ddAP-containing probes, 0.2 U/µL Tth Endonuclease IV (NEB), and 1 µL of target in 1X NEBuffer r2.1, with a total reaction volume of 10 µL for each sample. After incubation at 55°C for 30 minutes, 90 µL of lateral flow running buffer was added, then the lateral flow strip was added and allowed to develop for 5 minutes. Lateral flow strips were imaged using a GelDoc Go Gel Imaging System (BioRad) using the colorimetric mode and an exposure time of 0.15 seconds.

### qPCR Determination of *EGFR* Mutational Status

Lung cancer genomic extracts were purchased from BioChain. 93 lung cancer genomic extracts were classified using qPCR. T790M status was determined using the TaqMan SNP Genotyping Assay (C_156046749_20) with the TaqPath ProAmp Master Mix. Each 10-µL reaction contained 1X SNP Genotyping Assay, 1X TaqPath ProAmp Master Mix, and 1 µL of 100 ng/µL genomic DNA extract. A StepOnePlus Real-Time PCR instrument was used in genotyping mode with the following settings: Pre-Read, 60°C 30 seconds, Initial denature, 95°C 5 minutes, 60 cycles of Denature 95°C 15 seconds, Anneal/Extend/Read 60°C 60 seconds; Post-Read 60°C 30 seconds. Exon 19 deletion status was determined using custom primers and probes (Supplementary Table 5) with TaqPath ProAmp Master Mix. Each 10 µL reaction contained 400 nM each forward and reverse primer, 200 nM each probe, 1X TaqPath ProAmp Master Mix and 1 µL of 100 ng/µL genomic DNA extract. When plotting ΔRn of each channel against each other, clear clusters corresponding to wild-type *EGFR* were observed. For T790M mutant-containing samples had a noticeably high ΔRn on T790M probe, while Ex19del mutant-containing samples had noticeably low ΔRn on the Ex19del Probe.

### Four-plex TIMBER *EGFR* Mutational Analysis

Six lung cancer genomic samples with known *EGFR* mutations and six wild-type samples were amplified and rendered single stranded as previously described for RT-PCR with the following modifications, the reverse transcription step was removed, and only 30 cycles were used. TIMBER reactions were prepared using 100 nM of each probe, and 1 µL of PCR sample was added to a 40-µL reaction.

### Clinical Virus Sample Collection and Processing

A panel of 64 saliva samples, comprising 32 confirmed SARS-CoV-2 negative and 32 SARS-CoV-2 positive with Cycle threshold (Ct) values ranging from 17 to 23 were tested. Saliva/viral mRNA was extracted through a 2-minute heat treatment at 98°C, and the resulting lysate was diluted 1:4. Subsequently, target amplification was performed via a 35-minute RT-LAMP reaction at 65°C. The 64 de-identified saliva samples used in this study were obtained from the Biodesign Institute Clinical Testing Laboratory (Arizona State University). The specimens were collected for SARS-CoV-2 diagnostic purposes with the consent of the patients and provided as de-identified clinical remnants under the oversight of the Arizona State University Institutional Review Board (IRB ID: STUDY00011737). TIMBER reactions were prepared using 100 nM of probe, and 2 µL of RT-LAMP product was added to a 20-µL reaction.

### Fitting and Statistical Analysis

All data collected in triplicate. Error bars indicate standard deviation. Limit-of-detection of TIMBER was assessed using Graphpad Prism by fitting a 4PL curve to the data using a 1/Y^2^ weighting and back calculating the concentration that corresponded to the value 3 standard deviations above the mean fluorescence of the negative sample.

## Data Availability

All data produced in the present work are contained in the manuscript

## Author contributions

P.M.G. and A.A.G. conceived the idea. P.M.G. and Y.L. developed the methodology, designed, performed the experiments and data analysis. V.M. acquired the SARS-CoV-2 clinical samples. A.A.G. supervised the research and acquired funding. P.M.G., Y.L. and A.A.G. wrote the manuscript.

## Acknowledgments

This work was developed with funding from the Defense Advanced Research Projects Agency (DARPA), Contract No. N66001-23-2-4042; a National Institutes of Health (NIH) U01 award (1U01AI148319-01) and R01 award (1R01EB031893); and Boston University startup funds to A.A.G. This material is based upon work supported by the National Science Foundation Graduate Research Fellowship under Grant No. DGE-1840990 to P.M.G. The views, opinions and/or findings expressed are those of the authors and should not be interpreted as representing the official views or policies of the Department of Defense or the U.S. Government. The content is solely the responsibility of the authors and does not necessarily represent the official views of the National Institutes of Health. Any opinion, findings, and conclusions or recommendations expressed in this material are those of the authors(s) and do not necessarily reflect the views of the National Science Foundation.

