## Supplementary Figures for "Multiplexed Isothermal Nucleic Acid Detection Using Sequence-Specific Cleavage Mediated by Repair Endonucleases"

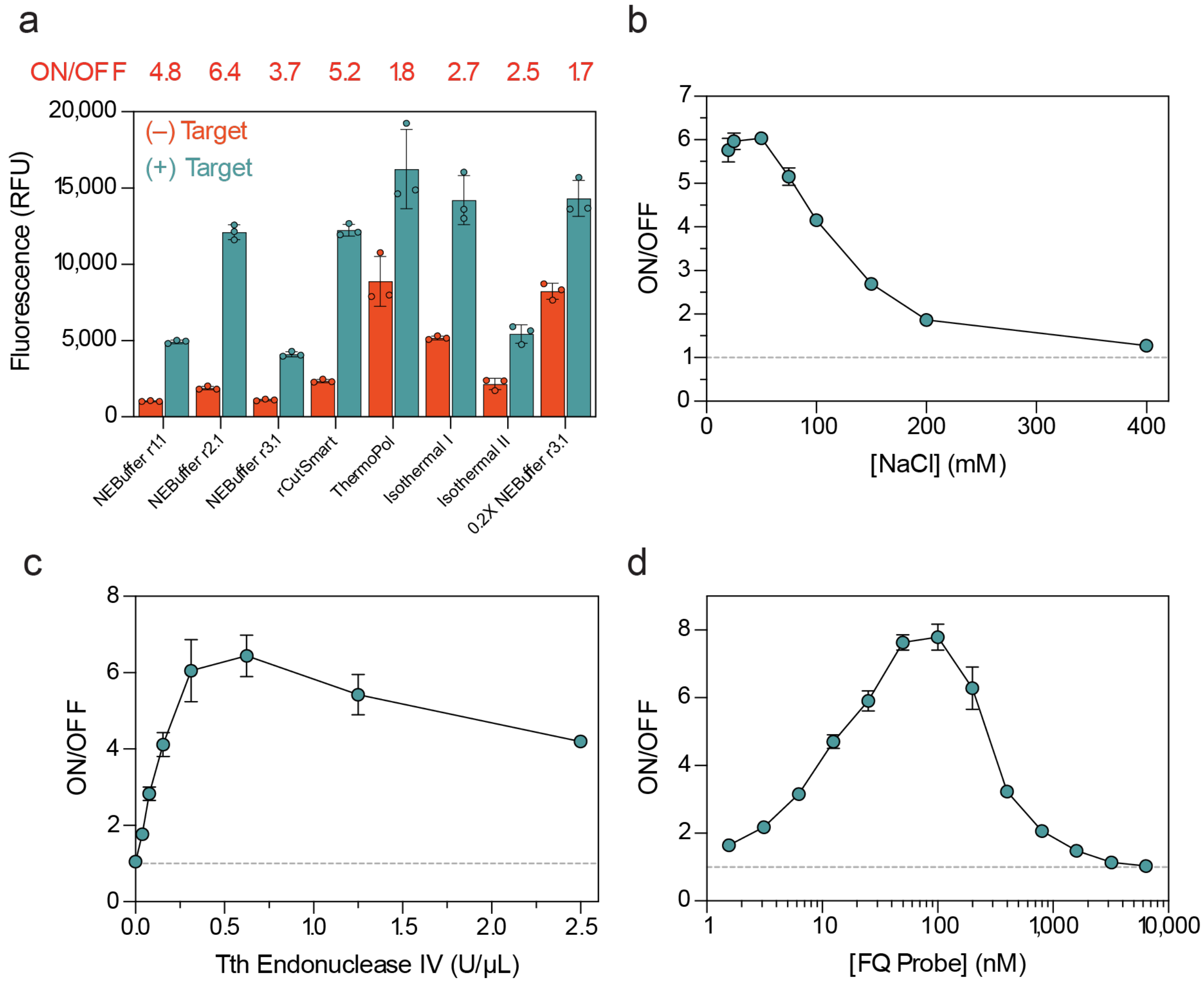


**Supplementary Figure 1. Optimization of TIMBER assay conditions. a,** Comparison of different assay buffer conditions for TIMBER. ON/OFF indicates the ratio of fluorescence intensity for samples containing 1 nM target vs an no-template control (NTC). **b,** Comparison of different NaCl concentrations on TIMBER performance. 10 mM Tris-HCl, 10 mM $MgCl_2$, 100 μg/ml recombinant albumin, pH 7.9 was supplemented with NaCl from 25 nM to 400 nM. **c,** Comparison of different concentrations of Tth endonuclease IV on TIMBER performance. **d,** Comparison between different FQ-Probe concentrations on TIMBER performance.

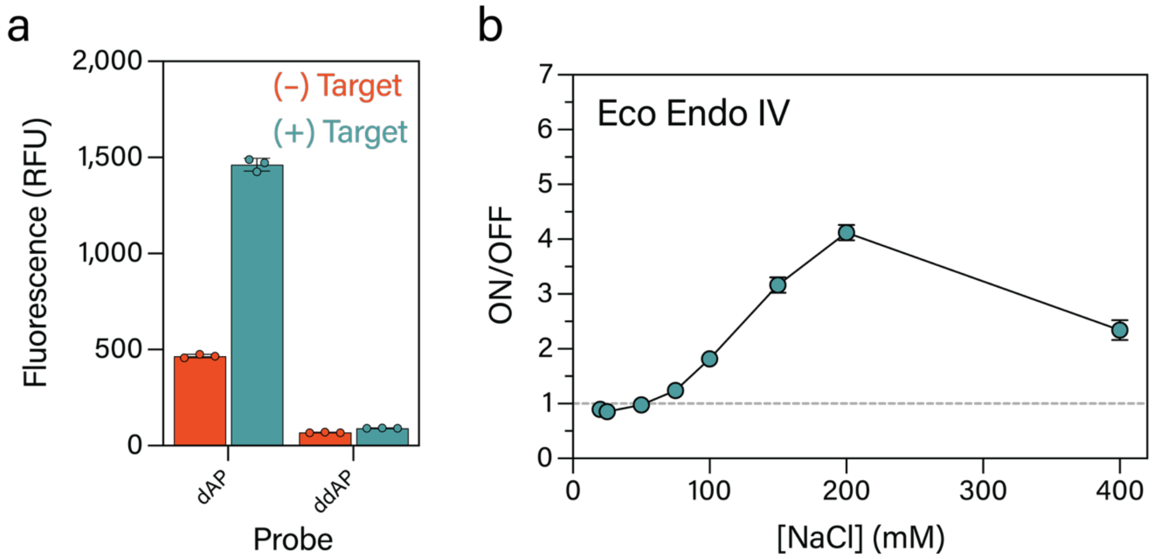


**Supplementary Figure 2. Mesothermal TIMBER using *E. coli* endonuclease III. a,** TIMBER using *E. coli* endonuclease III functions using a dAP containing probe at 37°C. **b,** NaCl concentration near 200 mM increases ON/OFF ratio when using ddAP substrate, indicating increased activity under moderate salt concentrations.

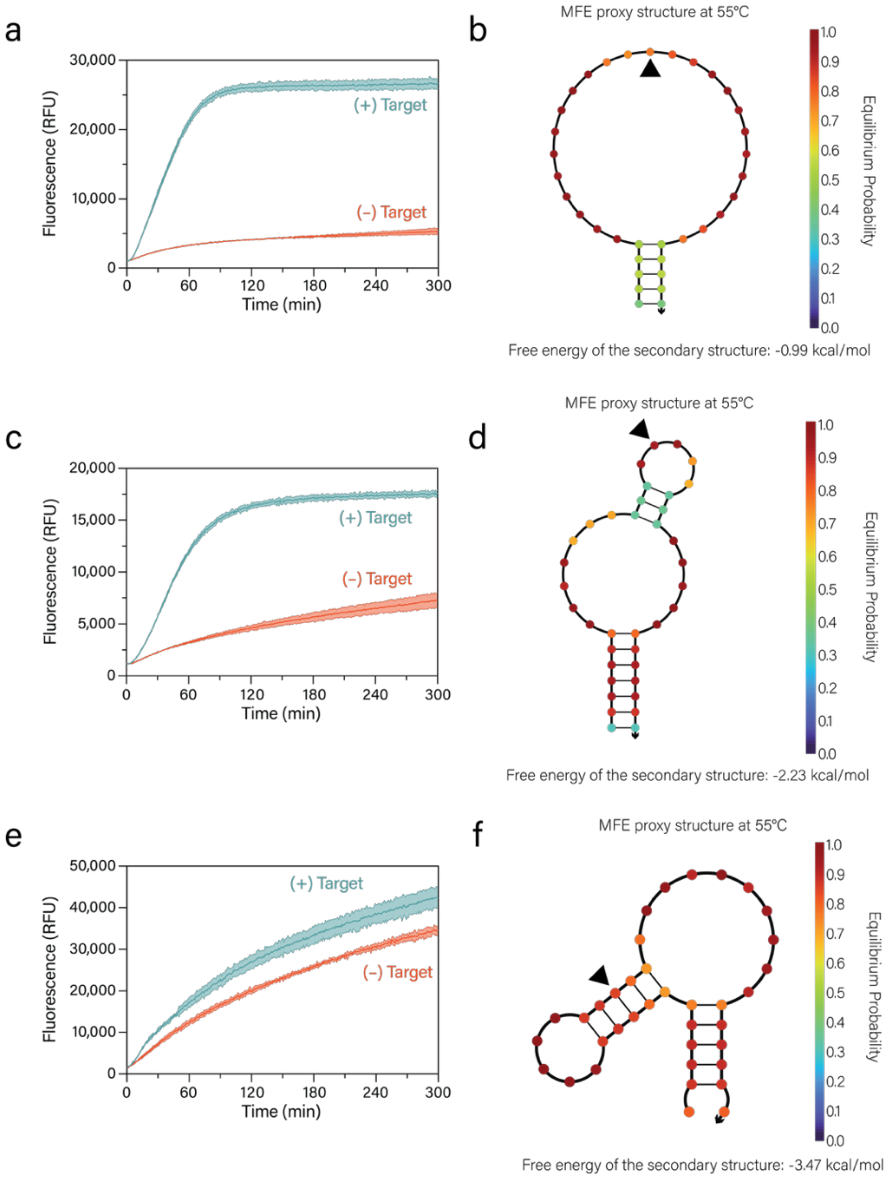


**Supplementary Figure 3. Sensitivity of TIMBER to probe secondary structure. a,** Response of TIMBER using ideal arbitrary probe in the presence and absence of 10 nM synthetic target. **b,** NUPACK predicted structure of this probe using the dna04 model, with 50 mM sodium and 10 mM magnesium, the abasic site was replaced with the original complementary base to the target for modeling purposes. Black triangle indicates the location of the abasic site on the probe. **c,** Response of TIMBER using *EGFR* T790M probe in the presence and absence of 10 nM synthetic Target. **d,** NUPACK predicted structure of *EGFR* T790M probe, indicating secondary structure, but with the abasic site located within a loop. **e,** Response of TIMBER using *EGFR* L858R probe in the presence and absence of 10 nM synthetic Target. **f,** NUPACK predicted structure for *EGFR* L858R probe, indicating that the secondary structure of the probe places the abasic site within a double stranded hairpin.

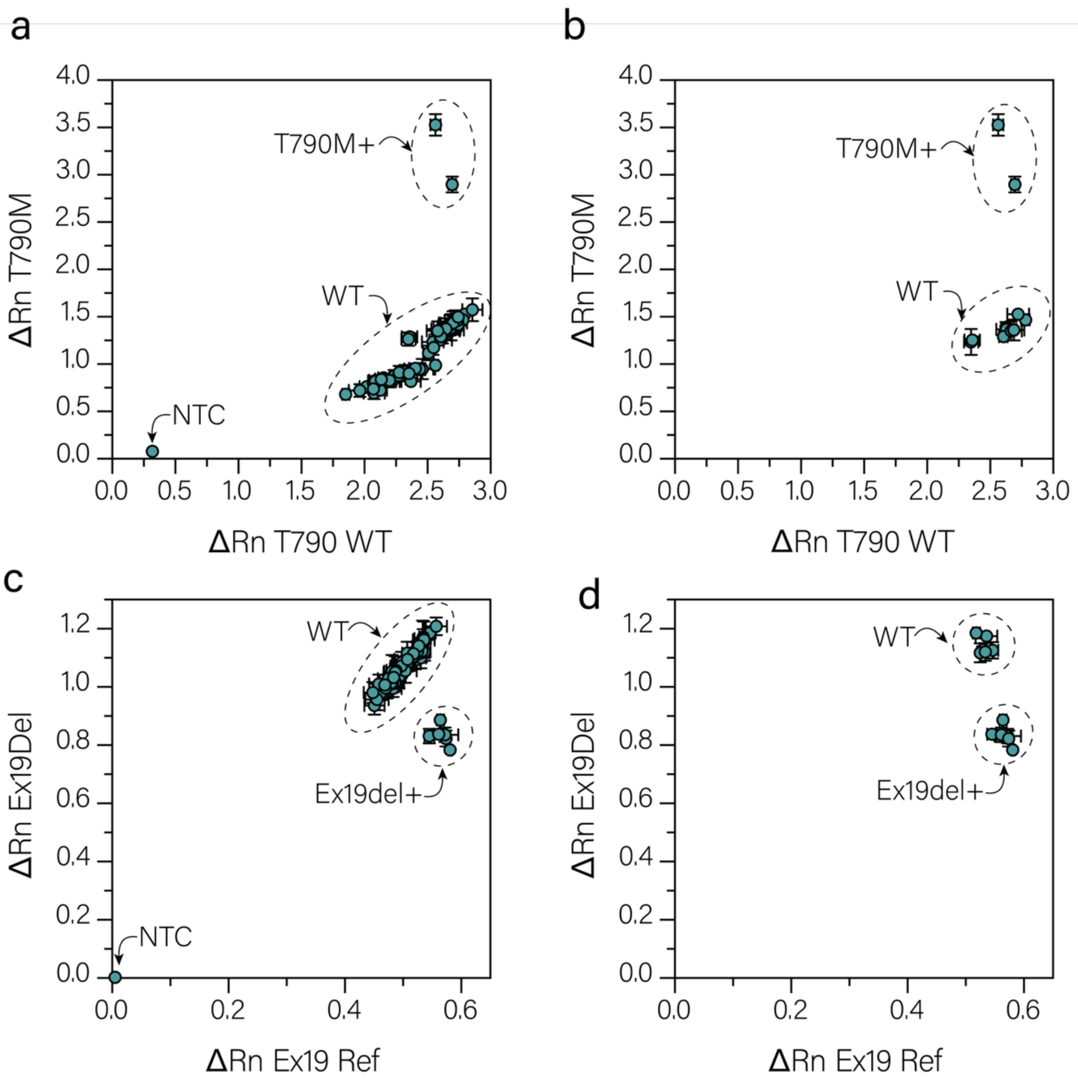


**Supplementary Figure 4. qPCR classification of lung cancer samples. a,** ΔRn for T790M qPCR probe vs. T790 WT probe for 62 lung cancer genomic DNA samples. WT samples appear as a clear cluster with low ΔRn for T790M, while two samples have high ΔRn for T790M indicating the presence of T790M mutation. **b,** ΔRn for T790M qPCR probe vs. T790 WT probe for down selected set of 12 samples used for TIMBER analysis. **c,** ΔRn for Ex19del qPCR probe vs. Ex19 Reference probe for 62 lung cancer genomic DNA samples. WT samples appear as a clear cluster along the diagonal, while six samples have low relative ΔRn for Ex19del indicating the presence of Ex19del mutation. **d,** ΔRn for Ex19del qPCR probe vs. Ex19 Reference probe for downselected set of 12 samples used for TIMBER analysis.

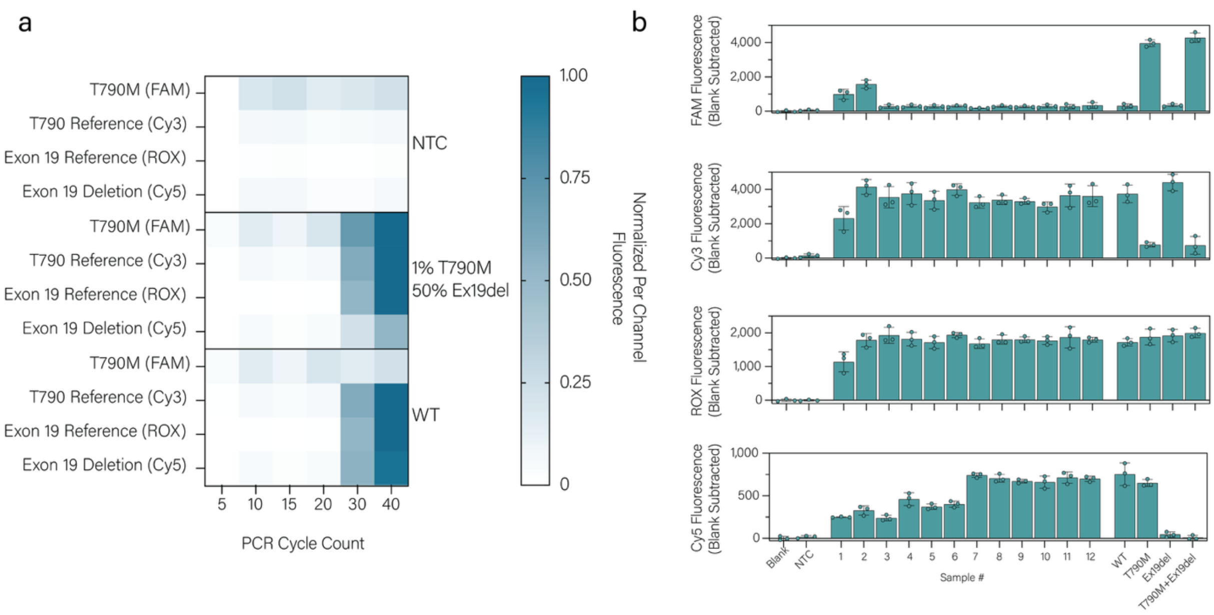


**Supplementary Figure 5. PCR-TIMBER for *EGFR* Mutational Analysis. a.** Optimization of PCR cycle count to enable mutational analysis via TIMBER. Simulated samples containing 1% T790M variant and 50% Ex19del variant, a simulated WT, and a NTC were examined at different cycle counts to ensure both low abundant T790M mutation and heterozygous Ex19del can be determined in the same sample. **b,** Fluorescence values for TIMBER mutational analysis of lung cancer samples.
